# Nocturnal hypertension and left ventricular diastolic dysfunction in diabetic patients with pre-heart failure phase: Prospective cohort HSCAA study

**DOI:** 10.1101/2023.07.17.23292802

**Authors:** Yonekazu Kidawara, Manabu Kadoya, Masataka Igeta, Akiko Morimoto, Akio Miyoshi, Miki Kakutani-Hatayama, Akinori Kanzaki, Kosuke Konishi, Yoshiki Kusunoki, Takashi Daimon, Masanori Asakura, Masaharu Ishihara, Hidenori Koyama

## Abstract

**Background:** Diabetes is an important risk factor of heart failure (HF) and is associated with left ventricular (LV) diastolic dysfunction. However, integrated importance of diabetes and its comorbid conditions, such as altered nocturnal blood pressure (BP) variation, as predictors of diastolic dysfunction is not known in pre-HF period. The present study was conducted as longitudinal examination of the predictive value of nocturnal hypertension profiles on progression of LV diastolic dysfunction in diabetic and non-diabetic patients without heart diseases.

**Methods:** Pre-heart failure 422 subjects (154 diabetes, 268 non-diabetes) were followed for 36.8 ± 18.2 months. The relationships among the patterns of nocturnal hypertension and the outcome of LV diastolic dysfunction, defined as increase in E/e’ >14, were investigated in the patients with and without diabetes.

**Results:** The interaction effect of the diabetes status and the patterns of nocturnal hypertension on the hazard rate of the occurrence of E/e’>14 was statistically significant (p=0.017). Kaplan-Meier analysis results revealed that diabetic patients with non-dipper (p=0.016 vs. dipper) and riser (p=0.007 vs. dipper) had a significantly greater risk for a diastolic dysfunction event. Furthermore, multivariable Cox proportional hazards analysis revealed that non-dipper (HR: 3.00; 95% CI: 1.11–8.06, p = 0.029) and riser (HR: 3.58; 95% CI:1.24–10.35, p = 0.018) patterns were significantly associated with elevated risk of the outcome of LV diastolic dysfunction. In contrast, no similar significant associations were found in non-diabetic patients.

**Conclusions:** During pre-HF periods, nocturnal hypertension is an important predictor for progression of LV diastolic dysfunction in diabetic patients.

**Non-standard Abbreviation and Acronyms:** HSCAA: Hyogo Sleep Cardio-Autonomic Atherosclerosis

## Introduction

Diabetes is one of the most common causes of heart failure (HF) and is associated with left ventricular (LV) diastolic dysfunction ^1–4^. Diabetes appears playing a fundamental role in the development of heart failure with preserved ejection fraction (HFpEF). In early stages, diabetic cardiomyopathy exhibits subclinical structural and functional abnormalities, including LV hypertrophy and fibrosis, and these pathophysiological changes are associated with subclinical diastolic dysfunction that often progresses to HFpEF ^5^. There is shown to be a strong association of prevalence between glycemic abnormalities and LV diastolic dysfunction ^6–8^. In addition to the hyperglycemic burden, the risk of HFpEF is shown to increase sharply with other risk factors including age, hypertension, and obesity ^9–11^. Nocturnal hypertension has been reported as a possible predictor of HFpEF ^12^. Of importance, another previous report had shown that nocturnal hypertension, such as non-dipper or riser pattern, is an important predictor of adverse events in patients with HFpEF ^13, 14^. Diabetic patients are reported to have higher nocturnal blood pressure (BP) and higher morning BP surge ^15^. Thus, mutual interactions among diabetes and nocturnal BP variations may potentially contribute to progression of diabetes cardiomyopathy, particularly diastolic dysfunction. However, it is unknown whether nocturnal hypertension contributes to the progression of LV diastolic dysfunction in pre-HF period in patients with or without diabetes.

This prospective investigation was conducted in association with the Hyogo Sleep Cardio-Autonomic Atherosclerosis (HSCAA) cohort study. Each impact of nocturnal hypertension patterns in pre-HF phase was examined as predictors of progression of LV diastolic dysfunction as compared with and without diabetes.

## Methods

### Study design and participants

The HSCAA study is an ongoing cohort investigation instituted as a review of cardio-metabolic risk factors, including sleep parameters, for elucidation of the clinical implications of cardiovascular and metabolic diseases ^16, 17^. From October 2010 to December 2018, 976 patients with at least one cardiovascular risk factor, including obesity, hypertension, dyslipidemia, diabetes mellitus, and chronic kidney disease, were enrolled. Patients (n=240) with ischemic heart disease, moderate to severe valvular heart disease, hypertrophic cardiomyopathy, atrial fibrillation, HF, or cardiac diastolic dysfunction [Transmitral early inflow velocity / Early diastolic tissue velocity (E/e’ >14)] according to the recommendations of the American Society of Echocardiography and the European Association of Cardiovascular Imaging ^18^ excluded. 226 patients with missing cardiac ultrasonography baseline data were also excluded. For the present analysis, the time of data cut-off was at December 31, 2019. As a result, 88 with a follow-up period <12 months were excluded. Finally, 422 (diabetes: n = 154, non-diabetes: n = 268) of those patients with a mean follow-up period of 36.8 ± 18.2 months were enrolled in the present study (**Figure 1**). Measurements of echocardiography were performed within 2 weeks of nocturnal BP assessment at the time of HSCAA study registration. Following the measurement of cardiac function at enrollment, follow-up echocardiography was planned to be performed each year as much as possible (mandatory at 1, 3 and 5 years) as previously described ^19^. The HSCAA study was approved by the institutional ethical committee of Hyogo College of Medicine (approval No. 2351) and informed written consent was obtained from each participant.

**Figure 1.**
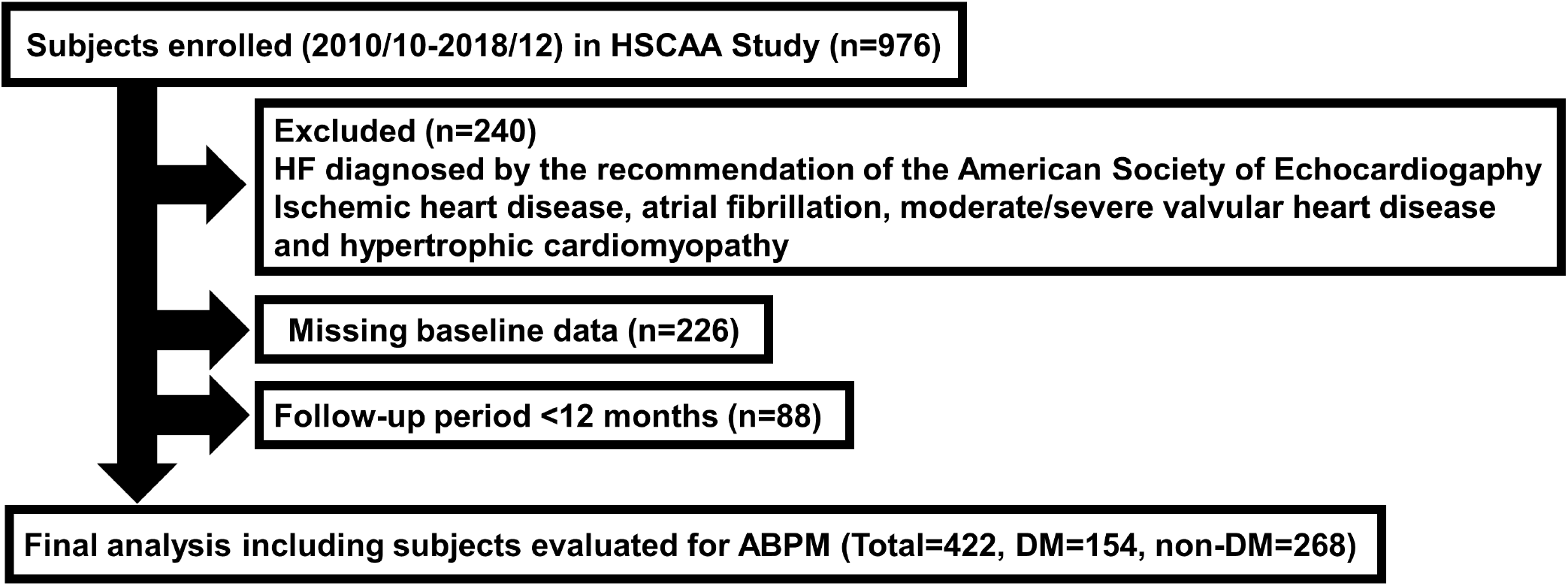
Flow of subject selection. HSCAA, Hyogo Sleep Cardio-Autonomic Atherosclerosis; HF, heart failure; ABPM, ambulatory blood pressure monitoring; DM, diabetes mellitus.

### Assessment of classic cardiovascular risk factors and determination of diabetes and dyslipidemia

We obtained the medical history of each subject, and measured height and body weight. Body mass index (BMI) was calculated as weight in kilograms divided by the square of height in meters (kg/m^2^). Smoking status was based on self-reported habit of cigarette smoking. Type 2 diabetes was diagnosed based on results showing fasting plasma glucose ≥126 mg/dl, causal plasma glucose ≥200 mg/dl, or 2-hour plasma glucose ≥200 mg/dl during a 75-g oral glucose tolerance test, or previous therapy for diabetes ^20^. Dyslipidemia was defined based on results showing low density lipoprotein (LDL) cholesterol (≥140 mg/dl), high density lipoprotein (HDL) cholesterol (≤40 mg/dl), elevated triglyceride level (≥150 mg/dl), or treatment for dyslipidemia ^21^. Blood samples were obtained in the morning after an overnight fast and then quickly centrifuged to obtain plasma. Whole blood was used for hemoglobin A1c, EDTA plasma for glucose and lipids, and serum for other biochemical assays. Glucose was measured using a glucose oxidase method. Serum creatinine concentration was determined by an enzymatic method. Estimated glomerular filtration rate (eGFR) in each subject was calculated using an equation for Japanese subjects, as follows: eGFR (ml/min/1.73 m^2^) = 194×age (years)^-0.287^×S-creatinine^-1.094^ (if female, ×0.739) ^22^.

### Ambulatory Blood Pressure Monitoring and determination of hypertension

Ambulatory Blood Pressure Monitoring (ABPM) was performed with a TM-2431 digital recorder and the obtained data were analyzed with a TM-9503 Doctor Pro3 software (A&D Co.Ltd., Tokyo, Japan), as previously described ^23–25^.

Measurements were made every 30 minutes over 48 hours (from evening to evening), resulting daytime and nighttime mean systolic blood pressure (SBP) and diastolic blood pressure (DBP) used as described ^25^. Basically, the latter 24 hour-recorded data were used for analyses. The major quality criteria used for an acceptable ABPM recording included the following: (1) minimum of 80% of the BP readings expected during the 24-h period; (2) no more than 2 nonconsecutive hours with <1 valid BP reading; and (3) no behaviors seriously affecting BP (afternoon nap, drinking and so on) as previously described ^23, 24^. Wake and sleep times were determined with the Actigraph (Ambulatory Monitoring, Inc., Ardley, New York, USA), as previously described ^26^. This device is worn on the wrist of the non-dominant arm and senses motion as acceleration, and then uses standard criteria to identify the onset and offset of sleep periods with a built-in algorithm. In this study, hypertension was defined as daytime SBP mean ≥140 mmHg or daytime DBP mean ≥90 mmHg evaluated by ABPM, or treatment for hypertension. The nocturnal SBP fall (%) was calculated as 100 × [1-sleep SBP/awake SBP ratio]. We classified the patients by the nocturnal SBP fall as follow: extreme-dippers if the fall was ≥ 20%, dippers if the fall was ≥ 10% but < 20%, non-dippers if it was ≥ 0% but < 10%, and risers if it was < 0% ^25^.

### Assessment of LV diastolic function

LV function was evaluated by echocardiography as previously described ^19, 27, 28^. Echocardiography examinations were performed with several different devices, including an IE-33, CX50 (Philips Healthcare, Massachusetts, USA), Artida, Aplio300 (Toshiba Medical System Co., Tochigi, Japan), and F75 (Hitachi-Aloka Medical, Tokyo, Japan), by six experienced sonographers and essentially repeated every one to two years to determine changes in LV function. To keep the consistency, the sonographers check the inter and intra examiner differences on every six months as regulated by standard operating procedure (SOP). The SOP was certified by the Japan Accreditation Board (International Organization for Standardization: ISO15189). The differences between the devices are also checked on every six months. The echocardiographic examinations were certified by the Japan Accreditation Board in 2016, and it was renewed in August 2020. LV diastolic function was also evaluated by echocardiography. LV ejection fraction (LVEF) was calculated using Teichholz’s formula in patients without LV segmental asynergy or deformation. In those with LV segmental asynergy or deformation, apical 2− and 4-chamber two-dimensional echo views and a modified version of Simpson’s method were used ^27^. Left ventricular mass index (LVMI) was calculated as follows: LVMI = 1.04 × (LVDd + IVST + RWT)^3^ - (LVDd)^3^ - 13.6 / 1000 / BSA, in which RWT (relative wall thickness) = IVST + PWT/LVDd, BSA (body surface area) = height (cm)^0.725^ × weight (kg)^0.425^ × 0.7184 (LVDd: left ventricular diastolic dimension, IVST: intraventricular septal thickness, PWT: posterior wall thickness). Transmitral early inflow velocity (E-wave), late diastolic filling velocity (A-wave), and E-wave deceleration time (DcT) were determined using pulsed Doppler echocardiography ^18^. Early diastolic tissue velocity (é) was measured in the septal basal region using tissue Doppler imaging, then the E/e′ ratio was calculated to obtain estimated LV filling pressure. Left arterial (LA) volume was assessed using Simpson’s biplane method and left atrial volume index (LAVI) was calculated by dividing LA volume by body surface area. Among these parameters, the primary outcome was the time to the LV diastolic dysfunction, which was defined as the time from registration into this study until the first development of E/e’>14 diagnosed at 1, 2, 3, 4 and 5 years. The E/e’>14 was previously recommended, as the value of 14 proved to be a good cutoff and is widely used in the world ^18^. If the outcome had not occurred, the follow up was censored at the earliest of either the case of loss-to-follow-up or the data locked date (December 31, 2019). Additionally, %change in E/e’ per year [100 × (last follow-up E/e’ - Baseline E/e’) / Baseline E/e’ / follow-up years] was also calculated to consider the annual effect as previously described ^19^.

### Statistical analyses

To compare mean and median values of the continuous variables of the baseline clinical patients’ characteristics between diabetes status (diabetic and non-diabetic population) and the dipping patterns, Welch’s t-test or Mann-Whitney U-test were used. For categorical variables, Fisher’s exact test was used. Since mutual interactions among diabetes and nocturnal BP variations may potentially contribute to progression of the LV diastolic dysfunction, the interaction effects of the diabetes status and the patterns of nocturnal hypertension on the time to E/e’>14 was evaluated on the Cox proportional hazards regression model. The covariates in each multivariable Cox proportional hazards analysis included age, gender, BMI, smoking history, presence of hypertension and dyslipidemia, HbA1c and baseline E/e’. The risk of HFpEF increases sharply with age ^9^, while female ^29^, hypertension ^10^, glucose abnormalities ^30^ and obesity ^11^ are also well recognized risk factors. In addition, risk factors for atherosclerosis, such as smoking and dyslipidemia, were also thought to influence myocardial function. Time to the LV diastolic dysfunction, E/e’>14, was compared among the groups of the dipping patterns using Kaplan-Meier and Cox proportional hazards regression analyses by the diabetes status. Since higher baseline E/e’ could be strongly associated with outcome, this was also included in the multivariable model. The Numbers at risk at each point for Kaplan-Meier analyses include subjects except those with the outcome prior to the point, lost-to-follow-up beyond the first year, or the data locked date been passed. The associations between %Change in E/e’ per year with nocturnal SBP fall were analyzed using multiple linear regression models. In comparisons among the dipping patterns, the extreme-dipper pattern was excluded because only one event of the primary outcome was observed in the pattern. All statistical analyses were performed with EZR, version 1.52 (Saitama Medical Center, Jichi Medical University, Saitama, Japan), a graphical user interface for R, version 4.02 (The R Foundation for Statistical Computing, Vienna, Austria) ^31^, and SAS software version 9.4 (SAS Institute Inc., Cary, NC, USA). All reported p-values are two-sided and were considered to be statistically significant when <0.05 without adjustment of multiplicity.

## Results

### Basal clinical characteristics in subjects with or without diabetes

The baseline characteristics of variables after dividing into those with and without diabetes are shown in **Table 1**. The distribution of dipping patterns in the patients with diabetes were as follows: risers, 20.1%; non-dippers, 39.7%; dippers, 35.1%; and extreme-dippers, 5.1%. Subjects with diabetes exhibited higher age and BMI, higher levels of systolic and diastolic BP, fasting plasma glucose, and HbA1c, lower levels of LDL and HDL-cholesterol, and higher prevalence of male gender, current smoking, and dyslipidemia. The use of angiotensin converting enzyme (ACE) inhibitor or angiotensin II receptor blocker (ARB) was significantly higher in the diabetes. Baseline A wave, E/A, e’ and E/e’, but not LVEF, were significantly worse in those with diabetes. %Change in E/e’ per year tended to be higher. Additionally, future diastolic dysfunction (E/e’ >14) was more frequently observed in those with diabetes than those without (23.3% vs. 11.1%, p=0.001). **Table 2** and **3** compare the basal clinical characteristics of the enrolled subjects with or without diabetes classified by nocturnal hypertension patterns. There were no clinically significant differences in age, gender, BMI, smoking, eGFR, HbA1c and medical hypertension or dyslipidemia treatment between non-dippers or risers with dippers in diabatic subjects. Baseline E/A, but not E/e’, was significantly different among non-dippers to dippers. %change of E/e’ per year was significantly higher in the risers (P=0.019). Whereas, there were no significant differences in baseline E/e’ and %change of E/e’ per year with non-diabetic subjects among 3 groups.

**Table 1.** Comparisons of baseline clinical characteristics.

| Variables | DM | Non-DM | P |
| --- | --- | --- | --- |
| Number | 154 | 268 |  |
| Age, years | 62.0 ± 10.7 | 57.4 ± 13.4 | <0.001 |
| Male gender, n (%) | 97 (62.9) | 116 (43.2) | <0.001 |
| Body mass index, kg/m<sup>2</sup> | 25.3 ± 4.7 | 23.7 ± 4.7 | <0.001 |
| Current smoking, n (%) | 51 (33.1) | 57 (21.2) | 0.010 |
| Systolic BP, mmHg | 127.1 ± 15.2 | 123.5 ± 15.4 | 0.022 |
| Diastolic BP, mmHg | 75.2 ± 8.4 | 75.3 ± 9.8 | 0.876 |
| LDL-cholesterol, mg/dL | 103.4 ± 32.8 | 119.3 ± 36.1 | <0.001 |
| HDL-cholesterol, mg/dL | 51.4 ± 15.1 | 57.9 ± 18.2 | <0.001 |
| Triacylglycerol, mg/dL | 122 (92-162) | 105 (80-153) | 0.012 |
| Blood urea nitrogen, mg/dL | 15.0 ± 4.9 | 14.1 ± 4.8 | 0.105 |
| Creatinine, mg/dL | 0.70 (0.56-0.81) | 0.66 (0.54-0.80) | 0.311 |
| eGFR, ml/min/1.73m<sup>2</sup> | 84.1 ± 24.7 | 83.5 ± 23.5 | 0.803 |
| Fasting plasma glucose, mg/dL | 125 (107-151) | 93 (87-99) | <0.001 |
| HbA1c (%) | 7.0 (6.3-7.9) | 5.3 (5.0-5.5) | <0.001 |
| Comorbidities |  |  |  |
| Hypertension, n (%) | 107 (69.4) | 166 (61.9) | 0.139 |
| Dyslipidemia, n (%) | 107 (69.4) | 144 (53.7) | 0.001 |
| Anti-hypertensive drugs |  |  |  |
| Calcium-channel blockers, n (%) | 64 (41.5) | 106 (39.5) | 0.757 |
| ACE inhibitor or ARB, n (%) | 63 (40.9) | 45 (16.7) | <0.001 |
| α or β blocker, n (%) | 15 (9.7) | 34 (12.6) | 0.431 |
| Diuretic agent, n (%) | 8 (5.1) | 10 (3.7) | 0.465 |
| Statin, n (%) | 68 (44.1) | 54 (20.1) | <0.001 |
| Anti-diabetic drugs |  |  |  |
| DPP-4 inhibitor, n (%) | 29 (18.8) | - | N/A |
| GLP-1 analogue, n (%) | 2 (1.2) | - | N/A |
| Sulfonylurea, n (%) | 27 (17.5) | - | N/A |
| Thiazolidine, n (%) | 5 (3.8) | - | N/A |
| Metformin, n (%) | 34 (22.0) | - | N/A |
| SGLT2 inhibitor, n (%) | 2 (1.2) | - | N/A |
| Insulin, n (%) | 18 (11.6) | - | N/A |
| Diabetes duration, years | 9.4 ± 8.7 | - | N/A |
| Diabetic microvascular complications |  |  |  |
| Neuropathy, n (%) | 57 (37.0) | - | N/A |
| Retinopathy, n (%) | 18 (11.6) | - | N/A |
| Nephropathy, n (%) | 34 (22.0) | - | N/A |
| LV function |  |  |  |
| LVMl | 99.6 ± 20.5 | 96.1 ± 22.1 | 0.110 |
| LVEF, % | 67.8 ± 6.3 | 70.0 ± 8.4 | 0.245 |
| E, cm/s | 59.7 ± 15.0 | 62.4 ± 15.8 | 0.095 |
| A, cm/s | 73.0 ± 14.9 | 66.9 ± 16.1 | <0.001 |
| E/A | 0.81 ± 0.24 | 0.99 ± 0.40 | <0.001 |
| DcT | 210 (175-242) | 200 (175-232) | 0.071 |
| e' | 6.2 ± 1.5 | 7.2 ± 2.0 | <0.001 |
| E/e' | 9.7 ± 2.2 | 8.9 ± 2.1 | <0.001 |
| LAVI | 28.7 ± 8.4 | 27.5 ± 7.7 | 0.327 |
| Future LV diastolic dysfunction, n (%) | 36 (23.3) | 30 (11.1) | 0.001 |
| %change of E/e' per year | 8.6 ± 22.4 | 5.5 ± 21.2 | 0.171 |
| Nocturnal SBP fall (%) | 7.3 ± 8.3 | 8.3 ± 10.1 | 0.262 |
| Patterns of nocturnal hypertension |  |  | 0.739 |
| Extreme-dipper, n (%) | 8 (5.1) | 14 (5.2) |  |
| Dipper, n (%) | 54 (35.1) | 103 (38.5) |  |
| Non-dipper, n (%) | 61 (39.7) | 108 (40.2) |  |
| Riser, n (%) | 31 (20.1) | 43 (16.1) |  |

Data are presented as the mean ± standard deviation or median (25th-75th percentile) for continuous variables or number (%) for dichotomous variables. P values are shown for comparisons of location parameters such as the means or median of two groups (two-sample Welch's t-test, Mann-Whitney U-test) or percentages (Fisher's exact test). CVD, cardiovascular disease; BP, blood pressure; LDL, low density lipoprotein; HDL, high density lipoprotein; eGFR, estimated glomerular filtration rate; ACE, angiotensin converting enzyme; ARB, angiotensin II receptor blocker; DPP, dipeptidyl peptidase; GLP, glucagon-like peptide.

**Table 2.** Comparisons of baseline clinical characteristics among nocturnal SBP fall in diabetic patients.

| Variables | Dipper | Non-dipper | Riser | P<br>(Non-dipper vs.<br>Dipper) | P<br>(Riser vs.<br>Dipper) |
| --- | --- | --- | --- | --- | --- |
| Number | 54 | 61 | 31 | - | - |
| Age, years | 62.2 ± 10.0 | 60.3 ± 11.2 | 63.4 ± 11.3 | 0.354 | 0.618 |
| Male gender, n (%) | 39 (72.2) | 36 (59.0) | 16 (51.6) | 0.171 | 0.063 |
| Body mass index, kg/m<sup>2</sup> | 24.7 ± 3.2 | 25.7 ± 4.6 | 24.9 ± 5.7 | 0.147 | 0.856 |
| Current smoking, n (%) | 18 (33.3) | 19 (31.1) | 10 (32.3) | 0.843 | 1.000 |
| Systolic BP, mmHg | 123.3 ± 12.9 | 128.1 ± 16.2 | 132.4 ± 16.3 | 0.079 | 0.010 |
| Diastolic BP, mmHg | 73.7 ± 7.7 | 76.7 ± 8.9 | 75.4 ± 8.9 | 0.054 | 0.376 |
| LDL-cholesterol, mg/dL | 95.6 ± 30.0 | 113.3 ± 33.9 | 102.9 ± 30.5 | 0.033 | 0.442 |
| HDL-cholesterol, mg/dL | 52.7 ± 17.4 | 50.4 ± 12.8 | 51.6 ± 14.6 | 0.428 | 0.752 |
| Triacylglycerol, mg/dL | 118 (85-149) | 131 (97-183) | 119.5 (88.5-160.5) | 0.091 | 0.616 |
| Blood urea nitrogen, mg/dL | 14.5 ± 3.7 | 14.3 ± 3.2 | 16.7 ± 7.6 | 0.825 | 0.196 |
| Creatinine, mg/dL | 0.70 (0.61-0.80) | 0.66 (0.54-0.80) | 0.67 (0.53-0.84) | 0.401 | 0.739 |
| eGFR, ml/min/1.73m<sup>2</sup> | 83.6 ± 19.8 | 87.3 ± 23.9 | 79.6 ± 31.6 | 0.379 | 0.541 |
| Fasting plasma glucose, mg/dL | 123 (108-139) | 131 (111-168) | 120 (103-156) | 0.088 | 0.713 |
| HbA1c (%) | 6.8 (6.3-7.5) | 7.4 (6.3-8.4) | 7.0 (6.6-8.0) | 0.108 | 0.180 |
| Comorbidities |  |  |  |  |  |
| Hypertension, n (%) | 33 (61.1) | 45 (73.8) | 24 (77.4) | 0.166 | 0.154 |
| Dyslipidemia, n (%) | 38 (70.4) | 42 (68.9) | 22 (71.0) | 1.000 | 1.000 |
| Anti-hypertensive drugs |  |  |  |  |  |
| Calcium-channel blockers, n (%) | 20 (37.0) | 30 (49.2) | 11 (35.5) | 0.258 | 1.000 |
| ACE inhibitor or ARB, n (%) | 20 (37.0) | 26 (42.6) | 14 (45.2) | 0.572 | 0.497 |
| $\alpha$ or $\beta$ blocker, n (%) | 2 (3.7) | 8 (13.1) | 4 (12.9) | 0.101 | 0.185 |
| Diuretic agent, n (%) | 3 (5.6) | 3 (4.9) | 2 (6.5) | 1.000 | 1.000 |
| Statin, n (%) | 27 (50.0) | 26 (42.6) | 11 (35.5) | 0.458 | 0.258 |
| Anti-diabetic drugs |  |  |  |  |  |
| DPP-4 inhibitor, n (%) | 9 (16.7) | 16 (26.2) | 4 (12.9) | 0.261 | 0.761 |
| GLP-1 analogue, n (%) | 1 (1.9) | 1 (1.6) | 0 (0.0) | 1.000 | 1.000 |
| Sulfonylurea, n (%) | 5 (9.3) | 14 (23.0) | 8 (25.8) | 0.076 | 0.060 |
| Thiazolidine, n (%) | 2 (3.7) | 2 (3.3) | 1 (6.5) | 1.000 | 0.620 |
| Metformin, n (%) | 12 (22.2) | 16 (26.2) | 5 (16.1) | 0.668 | 0.582 |
| SGLT2 inhibitor, n (%) | 0 (0.0) | 2 (3.3) | 0 (0.0) | 0.497 | 1.000 |
| Insulin, n (%) | 8 (14.8) | 6 (9.8) | 3 (9.7) | 0.569 | 0.739 |
| Diabetes duration, years | 8.2 ± 7.9 | 8.5 ± 7.9 | 14.7 ± 10.0 | 0.865 | <0.001 |
| Diabetic microvascular complications |  |  |  |  |  |
| Neuropathy, n (%) | 22 (40.7) | 18 (32.1) | 14 (51.9) | 0.429 | 0.356 |
| Retinopathy, n (%) | 8 (14.8) | 6 (9.8) | 3 (9.7) | 0.386 | 0.385 |
| Nephropathy, n (%) | 8 (14.7) | 14 (25.9) | 10 (38.4) | 0.300 | 0.020 |
| LV function |  |  |  |  |  |
| LVMI | 99.9 ± 20.5 | 97.8 ± 19.8 | 102.1 ± 23.3 | 0.591 | 0.668 |
| LVEF, % | 70.4 ± 4.2 | 66.1 ± 9.5 | 66.3 ± 4.1 | 0.203 | 0.044 |
| E, cm/s | 58.8 ± 16.1 | 62.8 ± 14.7 | 55.8 ± 13.6 | 0.177 | 0.369 |
| A, cm/s | 70.5 ± 11.6 | 73.1 ± 15.9 | 75.4 ± 16.5 | 0.339 | 0.170 |
| E/A | 0.79 ± 0.22 | 0.88 ± 0.28 | 0.73 ± 0.17 | 0.047 | 0.245 |
| DcT | 217 (175-247) | 208 (175-237) | 214 (181-238) | 0.529 | 0.889 |
| e' | 6.2 ± 1.6 | 6.4 ± 1.5 | 6.0 ± 1.2 | 0.455 | 0.637 |
| E/e' | 9.7 ± 2.1 | 10.0 ± 2.3 | 9.3 ± 2.1 | 0.542 | 0.379 |
| LAVI | 29.0 ± 9.3 | 28.8 ± 8.3 | 27.6 ± 7.1 | 0.927 | 0.603 |
| Future LV diastolic dysfunction, n (%) | 7 (13.0) | 19 (31.1) | 10 (32.3) | 0.025 | 0.047 |
| %change of E/e' per year | 4.4 ± 13.0 | 7.7 ± 20.1 | 20.2 ± 34.6 | 0.292 | 0.019 |
| Nocturnal SBP fall (%) | 14.0 ± 2.3 | 5.7 ± 2.4 | -5.0 ± 4.7 | <0.001 | <0.001 |

Data are presented as the mean ± standard deviation or median (25th-75th percentile) for continuous variables or number (%) for dichotomous variables. P values are shown for comparisons of location parameters such as the means or median of two groups (two-sample Welch's t-test, Mann-Whitney U-test) or percentages (Fisher's exact test). CVD, cardiovascular disease; BP, blood pressure; LDL, low density lipoprotein; HDL, high density lipoprotein; eGFR, estimated glomerular filtration rate; ACE, angiotensin converting enzyme; ARB, angiotensin II receptor blocker; DPP, dipeptidyl peptidase; GLP, glucagon-like peptide.

**Table 3.** Comparisons of baseline clinical characteristics among nocturnal SBP fall in non-diabetic patients.

| Variables | Dipper | Non-dipper | Riser | P<br>(Non-dipper<br>vs. Dipper) | P<br>(Riser vs.<br>Dipper) |
| --- | --- | --- | --- | --- | --- |
| Number | 103 | 108 | 43 | - | - |
| Age, years | 56.1 ± 13.5 | 57.7 ± 12.8 | 59.4 ± 15.8 | 0.377 | 0.199 |
| Male gender, n (%) | 41 (39.8) | 50 (46.2) | 19 (44.1) | 0.404 | 0.713 |
| Body mass index, kg/m<sup>2</sup> | 23.8 ± 4.0 | 24.1 ± 5.1 | 22.9 ± 5.6 | 0.581 | 0.312 |
| Current smoking, n (%) | 24 (23.3) | 21 (19.4) | 9 (20.9) | 0.507 | 0.831 |
| Systolic BP, mmHg | 121.7 ± 13.9 | 124.2 ± 14.8 | 127.3 ± 18.7 | 0.201 | 0.047 |
| Diastolic BP, mmHg | 74.4 ± 8.6 | 76.3 ± 10.1 | 76.0 ± 11.9 | 0.154 | 0.368 |
| LDL-cholesterol, mg/dL | 116.3 ± 32.3 | 119.7 ± 37.2 | 123.8 ± 43.7 | 0.610 | 0.396 |
| HDL-cholesterol, mg/dL | 56.8 ± 17.2 | 58.2 ± 17.8 | 56.5 ± 17.1 | 0.595 | 0.937 |
| Triacylglycerol, mg/dL | 103 (77-145) | 123 (90-173) | 93 (67-138) | 0.084 | 0.233 |
| Blood urea nitrogen, mg/dL | 13.7 ± 3.9 | 13.5 ± 4.6 | 16.1 ± 6.4 | 0.814 | 0.017 |
| Creatinine, mg/dL | 0.65 (0.54-0.82) | 0.64 (-.53-0.74) | 0.67 (0.57-0.80) | 0.615 | 0.557 |
| eGFR, ml/min/1.73m<sup>2</sup> | 84.8 ± 26.3 | 84.8 ± 20.7 | 80.0 ± 24.0 | 0.983 | 0.336 |
| Fasting plasma glucose, mg/dL | 92 (86-98) | 94 (88-99) | 94 (86-97) | 0.172 | 0.866 |
| HbA1c (%) | 5.2 (5.0-5.5) | 5.3 (5.0-5.5) | 5.3 (5.0-5.6) | 0.547 | 0.490 |
| Comorbidities |  |  |  |  |  |
| Hypertension, n (%) | 60 (58.3) | 67 (62.0) | 31 (72.1) | 0.673 | 0.136 |
| Dyslipidemia, n (%) | 53 (51.5) | 63 (58.3) | 23 (53.5) | 0.335 | 0.857 |
| Anti-hypertensive drugs |  |  |  |  |  |
| Calcium-channel blockers, n (%) | 45 (43.7) | 42 (38.9) | 16 (37.2) | 0.488 | 0.581 |
| ACE inhibitor or ARB, n (%) | 20 (19.4) | 16 (14.8) | 6 (14.0) | 0.465 | 0.487 |
| $\alpha$ or $\beta$ blocker, n (%) | 14 (13.6) | 12 (11.1) | 6 (14.0) | 0.677 | 1.000 |
| Diuretic agent, n (%) | 1 (1.0) | 7 (6.5) | 1 (2.3) | 0.065 | 0.504 |
| Statin, n (%) | 18 (17.5) | 24 (22.2) | 12 (27.9) | 0.491 | 0.180 |
| LV function |  |  |  |  |  |
| LVMI | 94.8 ± 18.5 | 96.4 ± 24.9 | 98.6 ± 19.6 | 0.621 | 0.273 |
| LVEF, % | 67.2 ± 11.8 | 70.8 ± 6.2 | 74.0 ± 2.1 | 0.305 | 0.288 |
| E, cm/s | 66.2 ± 15.8 | 59.4 ± 15.8 | 59.9 ± 13.1 | 0.002 | 0.015 |
| A, cm/s | 66.5 ± 15.6 | 67.0 ± 15.7 | 68.2 ± 18.6 | 0.838 | 0.593 |
| E/A | 1.04 ± 0.38 | 0.93 ± 0.32 | 0.98 ± 0.55 | 0.022 | 0.417 |
| DcT | 201 (172-229) | 200 (175-233) | 207 (179-224) | 0.948 | 0.701 |
| e' | 7.5 ± 2.0 | 7.09 ± 2.1 | 6.58 ± 2.0 | 0.119 | 0.010 |
| E/e' | 8.9 ± 2.1 | 8.8 ± 2.2 | 9.4 ± 1.9 | 0.739 | 0.139 |
| LAVI | 27.7 ± 7.1 | 27.4 ± 8.2 | 26.9 ± 8.3 | 0.878 | 0.674 |
| Future LV diastolic dysfunction, n (%) | 14 (13.6) | 11 (10.2) | 4 (9.3) | 0.525 | 0.588 |
| %change of E/e' per year | 6.7 ± 23.5 | 4.9 ± 21.5 | 5.9 ± 17.8 | 0.558 | 0.830 |
| Nocturnal SBP fall (%) | 13.9 ± 2.7 | 6.0 ± 2.5 | -5.7 ± 7.0 | <0.001 | <0.001 |

Data are presented as the mean ± standard deviation or median (25th-75th percentile) for continuous variables or number (%) for dichotomous variables. P values are shown for comparisons of location parameters such as the means or median of two groups (two-sample Welch's t-test, Mann-Whitney U-test) or percentages (Fisher's exact test). CVD, cardiovascular disease; BP, blood pressure; LDL, low density lipoprotein; HDL, high density lipoprotein; eGFR, estimated glomerular filtration rate; ACE, angiotensin converting enzyme; ARB, angiotensin II receptor blocker.

### Time-to-E/e’>14 analyses

The interaction effect of the diabetes status and the patterns of nocturnal hypertension on the hazard rate of the occurrence of E/e’>14 was statistically significant (p=0.017). Thus, the influence of the patterns of nocturnal hypertension on the occurrence of E/e’>14 was investigated for each diabetes status. As shown in **Figure 2**, Kaplan-Meier analysis findings indicated that subjects with non-dipper (log-rank test, p=0.016) and riser (p=0.007) pattern exhibited a significantly greater risk for decline in diastolic dysfunction as compared to the dipper pattern in diabetic patients, however, those with non-diabetic did not show significant risk for this outcome after matching. Note that only one primary outcome was observed in the extreme-dipper group, therefore, risk assessment analysis was not appropriate, and this group was excluded from the analysis.

**Figure 2.**
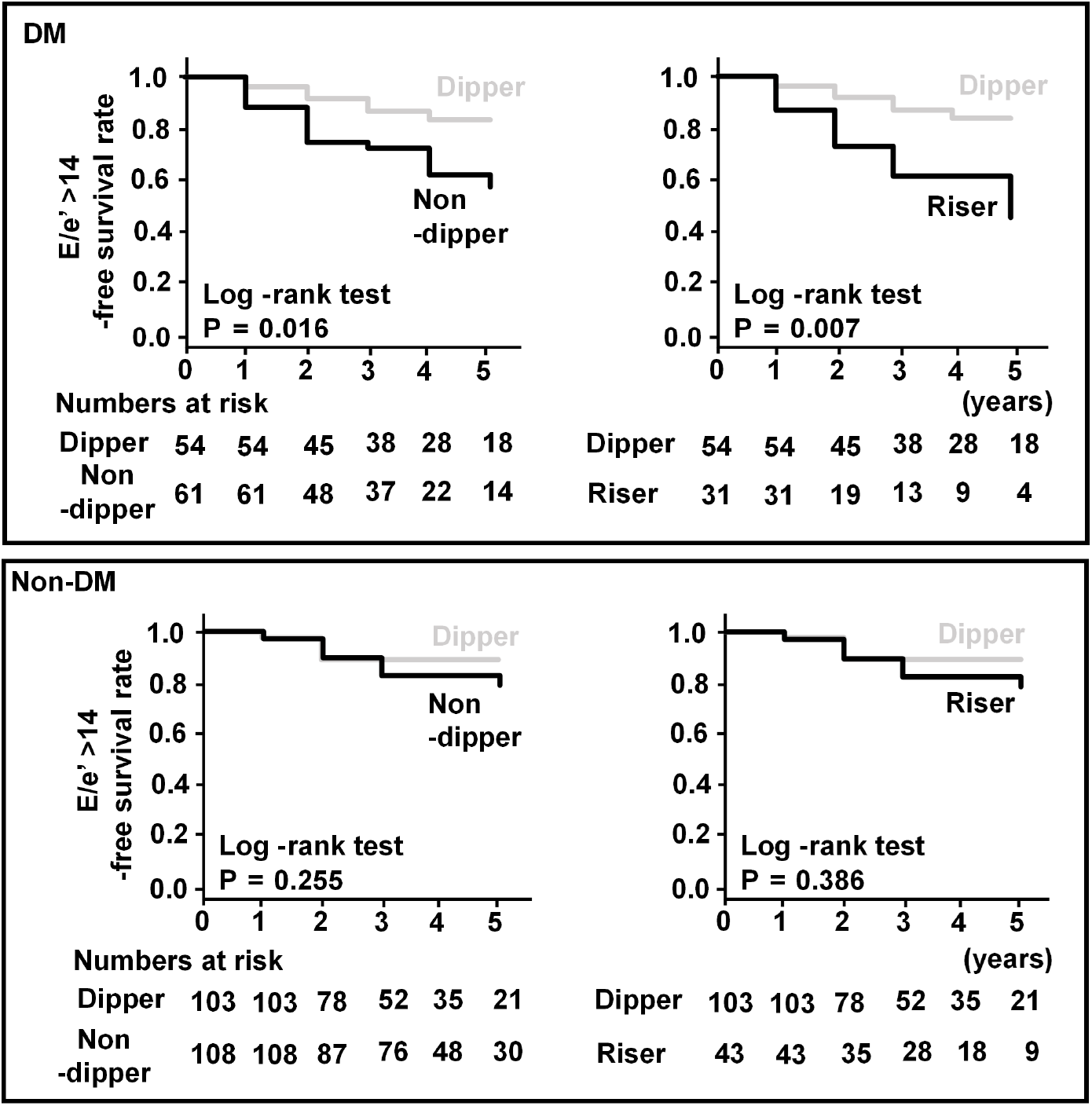
Kaplan-Meier analysis of the effects of nocturnal hypertension on LV diastolic dysfunction (defined by E/e’ >14) in the subjects with and without diabetes. Subjects in the diabetic and non-diabetic groups were compared dipper patterns with non-dipper or riser patterns. Probability was analyzed using a log-rank test.

To further evaluate the impact of nocturnal hypertension for future diastolic dysfunction, univariable and multivariable Cox proportional hazard regression analyses were performed (**Table 4**). In the patients with diabetes, non-dipper or riser pattern, and nocturnal SBP fall were significantly associated with future diastolic dysfunction in univariable analysis. As for multivariable analyses, non-dipper (Hazard ratio (HR): 3.00, 95% confidence interval (CI): 1.11-8.06, p=0.029) or riser (HR: 3.58, 95% CI: 1.24-10.35, p=0.018) pattern, and nocturnal SBP fall (HR: 0.94, 95% CI: 0.90-0.98, p=0.005) showed a significant association with the time to diastolic dysfunction, as defined by E/e’>14. However, in the patients with non-diabetes, nocturnal hypertension patterns and SBP fall were not significantly associated with diastolic dysfunction with either the univariable or multivariable Cox proportional hazards analysis.

**Table 4.**
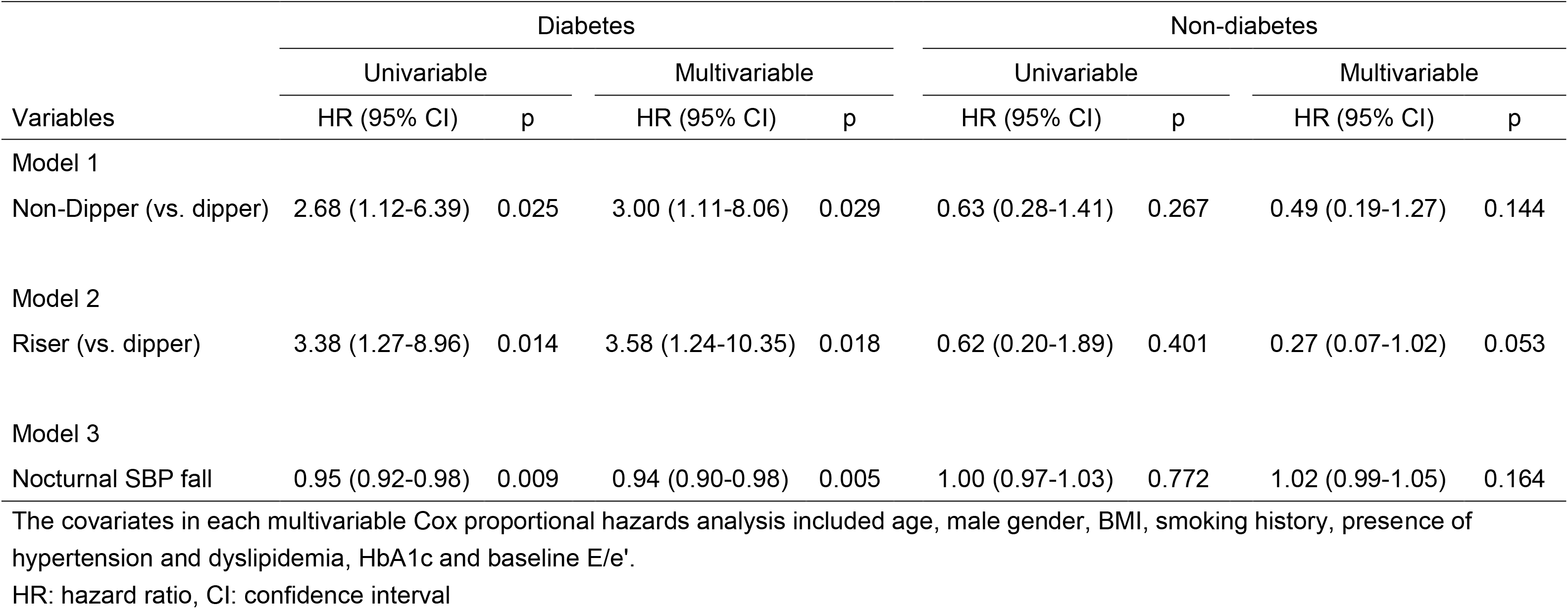
Univariable and multivariable Cox proportional hazards analyses of effects of nocturnal SBP fall on future cardiac diastolic dysfunction (E/e’>14) in the patients with and without diabetes.

| Variables | Diabetes |  |  |  | Non-diabetes |  |  |  |
| --- | --- | --- | --- | --- | --- | --- | --- | --- |
|  | Univariable |  | Multivariable |  | Univariable |  | Multivariable |  |
|  | HR (95% CI) | p | HR (95% CI) | p | HR (95% CI) | p | HR (95% CI) | p |
| Model 1 |  |  |  |  |  |  |  |  |
| Non-Dipper (vs. dipper) | 2.68 (1.12-6.39) | 0.025 | 3.00 (1.11-8.06) | 0.029 | 0.63 (0.28-1.41) | 0.267 | 0.49 (0.19-1.27) | 0.144 |
| Model 2 |  |  |  |  |  |  |  |  |
| Riser (vs. dipper) | 3.38 (1.27-8.96) | 0.014 | 3.58 (1.24-10.35) | 0.018 | 0.62 (0.20-1.89) | 0.401 | 0.27 (0.07-1.02) | 0.053 |
| Model 3 |  |  |  |  |  |  |  |  |
| Nocturnal SBP fall | 0.95 (0.92-0.98) | 0.009 | 0.94 (0.90-0.98) | 0.005 | 1.00 (0.97-1.03) | 0.772 | 1.02 (0.99-1.05) | 0.164 |

The covariates in each multivariable Cox proportional hazards analysis included age, male gender, BMI, smoking history, presence of hypertension and dyslipidemia, HbA1c and baseline E/e'.
HR: hazard ratio, CI: confidence interval

### Continuous E/e’ changes analyses

**Table 5** shows the results of multivariable linear regression analysis to examine the associations of nocturnal SBP fall with %changes of E/e’ per year, which were fully adjusted for clinical factors. In diabetic subjects, those results showed that riser pattern (β=15.54, p=0.006) and nocturnal SBP fall (β=-0.72, p<0.001) were still significantly associated with E/e’ changes adjusted for covariates. Associations of non-dipper pattern with changes in E/e’ did not reach statistically significant. However, in the patients with non-diabetes, nocturnal hypertension patterns and SBP fall were not significantly associated with %changes of E/e’ per year.

**Table 5.** Multiple linear regression analyses of nocturnal SBP fall for %change of E/e’ per year in the patients with and without diabetes.

| Variables | Diabetes |  | Non-diabetes |  |
| --- | --- | --- | --- | --- |
| | $\beta$ (SE) | p | $\beta$ (SE) | p |
| Model 1 |  |  |  |  |
| Non-Dipper (vs. dipper) | 5.29 (2.87) | 0.06 | -0.54 (2.05) | 0.792 |
| Model 2 |  |  |  |  |
| Riser (vs. dipper) | 15.54 (5.50) | 0.006 | 2.59 (2.58) | 0.317 |
| Model 3 |  |  |  |  |
| Nocturnal SBP fall | -0.72 (0.21) | <0.001 | -0.11 (0.08) | 0.186 |

\*The covariates in each multiple linear regression analysis models included age, male gender, BMI, smoking history, presence of hypertension, dyslipidemia, HbA1c and baseline E/e'.
SE: standard error

## Discussion

This is the first study to examine the impact of nocturnal hypertension on progression of cardiac diastolic dysfunction in diabetic and non-diabetic patients without heart failure. The greatest strength of this cohort study is that nocturnal hypertension is shown as an important predictor for progression of LV diastolic dysfunction in diabetic patients during pre-HF periods independent of other risk factors including age, obesity and glucose abnormalities. In contrast, nocturnal hypertension was not established as a significant risk factor for LV diastolic dysfunction in non-diabetic patients.

LV diastolic dysfunction is an important cause of HFpEF in individuals with diabetes and epidemiologic studies have shown the presence of diastolic dysfunction in type 2 diabetes patients ^32–34^. In our recent study, diabetic patients exhibited higher E/e’ than those with either normal or impaired glucose tolerance ^28^. In patients with Type 2 diabetes, E/e’ is shown to provide independent and incremental prognostic information predicting cardiovascular morbidity and mortality ^35^. Thus, the elevated E/e’ can be a good parameter of early diastolic impairment in diabetes, potentially foretelling diabetic cardiomyopathy. Longitudinal studies on diabetic patients are needed to better understand and predict this subtle, slow onset of diabetic cardiomyopathy. However, longitudinal studies evaluating changes in continuous parameters of LV diastolic function over time in subjects without clinically diagnosed HF are scant. In our study, frequent measurements of cardiac function have revealed a contributing factor to diabetic cardiomyopathy during pre-HF phase.

Potential mediators of associations between diabetes and cardiac diastolic dysfunction are not well clarified. A previous study showed that even in a population with normal glucose tolerance, a parameter of diastolic dysfunction was inversely associated with HbA1c ^36^. Variations of HbA1c may be a predictor of changes in E/e’ ^37^. Additionally, glycemic variability as measured by continuous glucose monitoring was also shown to be associated with left ventricular diastolic function in type 2 diabetes mellitus ^30^. However, in our current cohort, HbA1c was not a major predictor for an annual change in E/e’ (r=0.102, p=0.099). In population-based cohort studies, hyperinsulinemia at baseline predicted worsening of LV diastolic dysfunction. In our study, plasma insulin was not associated with annual changes of E/e’ in diabetic (r=0.029, p=0.753) and non-diabetic subjects (r=0.080, p=0.396). Another potential mediator includes presence of diabetic microangiopathy; a diastolic parameter E/e’ was shown to be worsened across groups of albuminuria severity ^38^. In our present cohort, however, presence of either type of diabetic microangiopathy (retinopathy, neuropathy, or nephropathy) was not significantly associated with annual changes in E/e’. Anti-diabetic agents may potentially affect progression of LV diastolic dysfunction, such as the sodium glucose cotransporter type 2 (SGLT2) inhibitors as recently shown to reduce worsening heart failure or death from cardiovascular causes in patients with heart failure ^39^. In our current study, only less than 2% of the patients were treated with SGLT2 inhibitors during registration period. Future surveys for anti-diabetic reagents during follow-up period may unveil their potential associations with progression of LV diastolic dysfunction.

Hypertension is also shown to be commonly associated with abnormalities in systolic function and LV diastolic filling ^40^. When compared to patients with HF with a reduced ejection fraction (HFrEF), patients with HFpEF are reported to more often have hypertension ^5^. ABPM has proven to be a more accurate to monitor and control BP. In a total of 150 outpatients treated with antihypertensive drugs for at least 1 year, the non-dipper status, controlled for age, gender and LVMI showed significant association with E/e’ ^41^. Similarly, a cross-sectional study included 376 hypertensive patients showed that most of the parameters of the LV diastolic function (including E/e’) were significantly deteriorated in the reverse dippers, with nocturnal systolic BP fall independently associated with LV diastolic function ^42^. Moreover, ABPM appears superior to conventional BP in association with 9.5-years later E/e′ in longitudinal study ^43^. However, only one longitudinal (over 20 years of follow up) study including 414 middle-aged subjects from OPERA cohort, 24-hour mean, daytime mean and nighttime mean pulse pressure, but neither different circadian BP profiles, showed significant association with the development of LV diastolic dysfunction as determined by E/e’ >=15, even after adjustment with significant baseline clinical characteristics ^44^. Moreover, diabetic patients were reported to have higher nocturnal blood pressure (BP) and higher morning BP surge ^15^, however, it was unknown whether nocturnal hypertension contributes to the progression of LV diastolic dysfunction in diabetic patients with pre-HF phase. Our current study is the first to focus on the predictive values of BP variation on LV diastolic dysfunction in diabetic as compared with non-diabetic subjects, and found that nocturnal hypertension was significantly associated with the event of E/e’ >14 and annual changes in E/e’ in diabetic, but not in non-diabetic subjects with pre-heart failure phase.

This study has several limitations. First, the findings of this study may not be generalized for all patients with different glycemic status. Second, we also have to interpret carefully the findings in the sub-group analysis of diabetes and non-diabetes, due to the limited numbers of events and the different distribution of the baseline clinical characteristics. Third, not all items of diagnostic criteria for cardiac diastolic dysfunction could not be evaluated, particularly atrial volume.

E/e’ has in several studies been found to lack accuracy in reflecting LV filling pressure in patients with preserved ejection fraction. E/e’ seemed to have better prognostic value in men compared to women, with E/ early diastolic strain rate (e’sr) may provide incremental information beyond that of E/e’ alone ^45^, which was not analyzed in this study. Nevertheless, our results provide important initial findings to unveil the pathophysiological mechanisms for HFpEF and diabetic cardiomyopathy.

In conclusion, during pre-HF periods, nocturnal hypertension is an important predictor for progression of LV diastolic dysfunction in diabetic patients.

## Data Availability

No data on this paper has been published.

## Acknowledgements

The authors are grateful for the excellent technical assistance of Tomoe Ushitani and Ai Matsumoto. We also wish to thank the other investigators and staff, as well as the participants of the Hyogo Sleep Cardio-Autonomic Atherosclerosis study for their valuable contributions.

## Supplemental Material

None

## References

1. Kannel WB, Hjortland M, Castelli WP. Role of diabetes in congestive heart failure: the Framingham study. Am J Cardiol. 1974;34:29–34. doi: 10.1016/0002-9149(74)90089-7

2. Aronow WS, Ahn C. Incidence of heart failure in 2,737 older persons with and without diabetes mellitus. Chest. 1999;115:867–868. doi: 10.1378/chest.115.3.867

3. Nichols GA, Gullion CM, Koro CE, Ephross SA, Brown JB. The incidence of congestive heart failure in type 2 diabetes: an update. Diabetes Care. 2004;27:1879–1884. doi: 10.2337/diacare.27.8.1879

4. Redfield MM, Jacobsen SJ, Burnett JC, Jr., Mahoney DW, Bailey KR, Rodeheffer RJ. Burden of systolic and diastolic ventricular dysfunction in the community: appreciating the scope of the heart failure epidemic. JAMA. 2003;289:194–202. doi: 10.1001/jama.289.2.194

5. Owan TE, Hodge DO, Herges RM, Jacobsen SJ, Roger VL, Redfield MM. Trends in prevalence and outcome of heart failure with preserved ejection fraction. N Engl J Med. 2006;355:251–259. doi: 10.1056/NEJMoa052256

6. Thrainsdottir IS, Aspelund T, Thorgeirsson G, Gudnason V, Hardarson T, Malmberg K, Sigurdsson G, Ryden L. The association between glucose abnormalities and heart failure in the population-based Reykjavik study. Diabetes Care. 2005;28:612–616. doi: 10.2337/diacare.28.3.612

7. Lind M, Bounias I, Olsson M, Gudbjornsdottir S, Svensson AM, Rosengren A. Glycaemic control and incidence of heart failure in 20,985 patients with type 1 diabetes: an observational study. Lancet. 2011;378:140–146. doi: 10.1016/S0140-6736(11)60471-6

8. Stratton IM, Adler AI, Neil HA, Matthews DR, Manley SE, Cull CA, Hadden D, Turner RC, Holman RR. Association of glycaemia with macrovascular and microvascular complications of type 2 diabetes (UKPDS 35): prospective observational study. BMJ. 2000;321:405–412. doi: 10.1136/bmj.321.7258.405

9. Borlaug BA, Redfield MM. Diastolic and systolic heart failure are distinct phenotypes within the heart failure spectrum. Circulation. 2011;123:2006–2013; discussion 2014. doi: 10.1161/CIRCULATIONAHA.110.954388

10. Dunlay SM, Roger VL, Weston SA, Bangerter LR, Killian JM, Griffin JM. Patient and Spousal Health and Outcomes in Heart Failure. Circ Heart Fail. 2017;10. doi: 10.1161/CIRCHEARTFAILURE.117.004088

11. Rao VN, Zhao D, Allison MA, Guallar E, Sharma K, Criqui MH, Cushman M, Blumenthal RS, Michos ED. Adiposity and Incident Heart Failure and its Subtypes: MESA (Multi-Ethnic Study of Atherosclerosis). JACC Heart Fail. 2018;6:999–1007. doi: 10.1016/j.jchf.2018.07.009

12. Komori T, Eguchi K, Saito T, Hoshide S, Kario K. Riser Pattern: Another Determinant of Heart Failure With Preserved Ejection Fraction. J Clin Hypertens (Greenwich). 2016;18:994–999. doi: 10.1111/jch.12818

13. Komori T, Eguchi K, Saito T, Hoshide S, Kario K. Riser Pattern Is a Novel Predictor of Adverse Events in Heart Failure Patients With Preserved Ejection Fraction. Circ J. 2017;81:220–226. doi: 10.1253/circj.CJ-16-0740

14. Ueda T, Kawakami R, Nakada Y, Nakano T, Nakagawa H, Matsui M, Nishida T, Onoue K, Soeda T, Okayama S, et al. Differences in blood pressure riser pattern in patients with acute heart failure with reduced mid-range and preserved ejection fraction. ESC Heart Fail. 2019;6:1057–1067. doi: 10.1002/ehf2.12500

15. Sun L, Yan B, Gao Y, Su D, Peng L, Jiao Y, Wang Y, Han D, Wang G. Relationship between blood pressure reverse dipping and type 2 diabetes in hypertensive patients. Sci Rep. 2016;6:25053. doi: 10.1038/srep25053

16. Kadoya M, Koyama H, Kurajoh M, Kanzaki A, Kakutani-Hatayama M, Okazaki H, Shoji T, Moriwaki Y, Yamamoto T, Emoto M, et al. Sleep, cardiac autonomic function, and carotid atherosclerosis in patients with cardiovascular risks: HSCAA study. Atherosclerosis. 2015;238:409–414. doi: 10.1016/j.atherosclerosis.2014.12.032

17. Kadoya M, Kurajoh M, Kakutani-Hatayama M, Morimoto A, Miyoshi A, Kosaka-Hamamoto K, Shoji T, Moriwaki Y, Inaba M, Koyama H. Low sleep quality is associated with progression of arterial stiffness in patients with cardiovascular risk factors: HSCAA study. Atherosclerosis. 2018;270:95–101. doi: 10.1016/j.atherosclerosis.2018.01.039

18. Nagueh SF, Smiseth OA, Appleton CP, Byrd BF, 3rd, Dokainish H, Edvardsen T, Flachskampf FA, Gillebert TC, Klein AL, Lancellotti P, et al. Recommendations for the Evaluation of Left Ventricular Diastolic Function by Echocardiography: An Update from the American Society of Echocardiography and the European Association of Cardiovascular Imaging. J Am Soc Echocardiogr. 2016;29:277–314. doi: 10.1016/j.echo.2016.01.011

19. Kidawara Y, Kadoya M, Morimoto A, Daimon T, Kakutani-Hatayama M, Kosaka-Hamamoto K, Miyoshi A, Konishi K, Kusunoki Y, Shoji T, et al. Sleep Apnea and Physical Movement During Sleep, But Not Sleep Duration, Are Independently Associated With Progression of Left Ventricular Diastolic Dysfunction: Prospective Hyogo Sleep Cardio-Autonomic Atherosclerosis Cohort Study. J Am Heart Assoc. 2022;11:e024948. doi: 10.1161/JAHA.121.024948

20. American Diabetes A. Diagnosis and classification of diabetes mellitus. Diabetes Care. 2004;27 Suppl 1:S5–S10.

21. Teramoto T, Sasaki J, Ishibashi S, Birou S, Daida H, Dohi S, Egusa G, Hiro T, Hirobe K, Iida M, et al. Executive summary of the Japan Atherosclerosis Society (JAS) guidelines for the diagnosis and prevention of atherosclerotic cardiovascular diseases in Japan-2012 version. J Atheroscler Thromb. 2013;20:517–523.

22. Matsuo S, Imai E, Horio M, Yasuda Y, Tomita K, Nitta K, Yamagata K, Tomino Y, Yokoyama H, Hishida A, et al. Revised equations for estimated GFR from serum creatinine in Japan. Am J Kidney Dis. 2009;53:982–992. doi: 10.1053/j.ajkd.2008.12.034

23. Kadoya M, Koyama H, Kanzaki A, Kurajoh M, Hatayama M, Shiraishi J, Okazaki H, Shoji T, Moriwaki Y, Yamamoto T, et al. Plasma brain-derived neurotrophic factor and reverse dipping pattern of nocturnal blood pressure in patients with cardiovascular risk factors. PLoS One. 2014;9:e105977. doi: 10.1371/journal.pone.0105977

24. Kadoya M, Koyama H, Kurajoh M, Naka M, Miyoshi A, Kanzaki A, Kakutani M, Shoji T, Moriwaki Y, Yamamoto T, et al. Associations of Sleep Quality and Awake Physical Activity with Fluctuations in Nocturnal Blood Pressure in Patients with Cardiovascular Risk Factors. PLoS One. 2016;11:e0155116. doi: 10.1371/journal.pone.0155116

25. Kario K, Pickering TG, Umeda Y, Hoshide S, Hoshide Y, Morinari M, Murata M, Kuroda T, Schwartz JE, Shimada K. Morning surge in blood pressure as a predictor of silent and clinical cerebrovascular disease in elderly hypertensives: a prospective study. Circulation. 2003;107:1401–1406.

26. Kadoya M, Koyama S, Morimoto A, Miyoshi A, Kakutani M, Hamamoto K, Kurajoh M, Shoji T, Moriwaki Y, Koshiba M, et al. Serum Macro TSH Level is Associated with Sleep Quality in Patients with Cardiovascular Risks - HSCAA Study. Sci Rep. 2017;7:44387. doi: 10.1038/srep44387

27. Okuhara Y, Asakura M, Orihara Y, Morisawa D, Matsumoto Y, Naito Y, Tsujino T, Ishihara M, Masuyama T. Reduction in Left Ventricular Ejection Fraction is Associated with Subsequent Cardiac Events in Outpatients with Chronic Heart Failure. Sci Rep. 2019;9:17271. doi: 10.1038/s41598-019-53697-y

28. Morimoto A, Kadoya M, Kakutani-Hatayama M, Kosaka-Hamamoto K, Miyoshi A, Shoji T, Goda A, Asakura M, Koyama H. Subclinical decrease in cardiac autonomic and diastolic function in patients with metabolic disorders: HSCAA study. Metabol Open. 2020;5:100025. doi: 10.1016/j.metop.2020.100025

29. Lam CSP, Chandramouli C. Fat, Female, Fatigued: Features of the Obese HFpEF Phenotype. JACC Heart Fail. 2018;6:710–713. doi: 10.1016/j.jchf.2018.06.006

30. Yokota S, Tanaka H, Mochizuki Y, Soga F, Yamashita K, Tanaka Y, Shono A, Suzuki M, Sumimoto K, Mukai J, et al. Association of glycemic variability with left ventricular diastolic function in type 2 diabetes mellitus. Cardiovasc Diabetol. 2019;18:166. doi: 10.1186/s12933-019-0971-5

31. Kanda Y. Investigation of the freely available easy-to-use software ‘EZR’ for medical statistics. Bone Marrow Transplant. 2013;48:452–458. doi: 10.1038/bmt.2012.244

32. Mosley JD, Levinson RT, Brittain EL, Gupta DK, Farber-Eger E, Shaffer CM, Denny JC, Roden DM, Wells QS. Clinical Features Associated With Nascent Left Ventricular Diastolic Dysfunction in a Population Aged 40 to 55 Years. Am J Cardiol. 2018;121:1552–1557. doi: 10.1016/j.amjcard.2018.02.042

33. Nayor M, Enserro DM, Xanthakis V, Larson MG, Benjamin EJ, Aragam J, Mitchell GF, Vasan RS. Comorbidities and Cardiometabolic Disease: Relationship With Longitudinal Changes in Diastolic Function. JACC Heart Fail. 2018;6:317–325. doi: 10.1016/j.jchf.2017.12.018

34. Reis JP, Allen NB, Bancks MP, Carr JJ, Lewis CE, Lima JA, Rana JS, Gidding SS, Schreiner PJ. Duration of Diabetes and Prediabetes During Adulthood and Subclinical Atherosclerosis and Cardiac Dysfunction in Middle Age: The CARDIA Study. Diabetes Care. 2018;41:731–738. doi: 10.2337/dc17-2233

35. Lassen MCH, Jensen MT, Biering-Sorensen T, Mogelvang R, Fritz-Hansen T, Vilsboll T, Rossing P, Jorgensen PG. Prognostic value of ratio of transmitral early filling velocity to early diastolic strain rate in patients with Type 2 diabetes. Eur Heart J Cardiovasc Imaging. 2019;20:1171–1178. doi: 10.1093/ehjci/jez075

36. Di Pino A, Mangiafico S, Urbano F, Scicali R, Scandura S, D’Agate V, Piro S, Tamburino C, Purrello F, Rabuazzo AM. HbA1c Identifies Subjects With Prediabetes and Subclinical Left Ventricular Diastolic Dysfunction. J Clin Endocrinol Metab. 2017;102:3756–3764. doi: 10.1210/jc.2017-00954

37. Li S, Zheng Z, Tang X, Zhong J, Liu X, Zhao Y, Chen L, Zhu J, Liu J, Chen Y. Impact of HbA1c variability on subclinical left ventricular remodeling and dysfunction in patients with type 2 diabetes mellitus. Clin Chim Acta. 2020;502:159–166. doi: 10.1016/j.cca.2019.12.006

38. Jorgensen PG, Biering-Sorensen T, Mogelvang R, Fritz-Hansen T, Vilsboll T, Rossing P, Jensen JS. Presence of micro- and macroalbuminuria and the association with cardiac mechanics in patients with type 2 diabetes. Eur Heart J Cardiovasc Imaging. 2018;19:1034–1041. doi: 10.1093/ehjci/jex231

39. McMurray JJV, Solomon SD, Inzucchi SE, Kober L, Kosiborod MN, Martinez FA, Ponikowski P, Sabatine MS, Anand IS, Belohlavek J, et al. Dapagliflozin in Patients with Heart Failure and Reduced Ejection Fraction. N Engl J Med. 2019;381:1995–2008. doi: 10.1056/NEJMoa1911303

40. Schillaci G, Pasqualini L, Verdecchia P, Vaudo G, Marchesi S, Porcellati C, de Simone G, Mannarino E. Prognostic significance of left ventricular diastolic dysfunction in essential hypertension. J Am Coll Cardiol. 2002;39:2005–2011. doi: 10.1016/s0735-1097(02)01896-x

41. Seo HS, Kang TS, Park S, Choi EY, Ko YG, Choi D, Ha J, Rim SJ, Chung N. Non-dippers are associated with adverse cardiac remodeling and dysfunction (R1). Int J Cardiol. 2006;112:171–177. doi: 10.1016/j.ijcard.2005.08.038

42. Ivanovic BA, Tadic MV, Celic VP. To dip or not to dip? The unique relationship between different blood pressure patterns and cardiac function and structure. J Hum Hypertens. 2013;27:62–70. doi: 10.1038/jhh.2011.83

43. Wei FF, Yang WY, Thijs L, Zhang ZY, Cauwenberghs N, Van Keer J, Huang QF, Mujaj B, Kuznetsova T, Allegaert K, et al. Conventional and Ambulatory Blood Pressure as Predictors of Diastolic Left Ventricular Function in a Flemish Population. J Am Heart Assoc. 2018;7. doi: 10.1161/JAHA.117.007868

44. Paakko TJW, Renko RJ, Perkiomaki JS, Kesaniemi YA, Ylitalo AS, Lumme JA, Huikuri HV, Ruskoaho H, Vuolteenaho O, Ukkola OH. Ambulatory Pulse Pressure Predicts the Development of Left Ventricular Diastolic Dysfunction in Over 20 Years of Follow-up. Am J Hypertens. 2017;30:985–992. doi: 10.1093/ajh/hpx087

45. Lassen MCH, Sengelov M, Qasim A, Jorgensen PG, Bruun NE, Olsen FJ, Fritz-Hansen T, Gislason G, Biering-Sorensen T. Ratio of Transmitral Early Filling Velocity to Early Diastolic Strain Rate Predicts All-Cause Mortality in Heart Failure with Reduced Ejection Fraction. J Card Fail. 2019;25:877–885. doi: 10.1016/j.cardfail.2019.07.007

